# Timing of cholecystectomy after percutaneous cholecystostomy for acute cholecystitis- A systematic review and meta-analysis

**DOI:** 10.1101/2021.06.06.21258426

**Authors:** Bhavin Vasavada, Hardik Patel

## Abstract

**Introduction:** There is a controversy about the optimum timing of cholecystectomy after percutaneous cholecystostomy.

This systematic review and meta-analysis aimed to evaluate outcomes of early versus late cholecystectomy after percutaneous cholecystostomy.

**Methods:** The study was conducted according to the Preferred Reporting Items for Systematic Reviews and Meta-Analyses (PRISMA) statement and MOOSE guidelines. Heterogeneity was measured using Q tests and I2 statistics. The random-effects model was used. We evaluated cholecystectomy performed at different periods after percutaneous cholecystostomy within 72 hours or later, within or after one week or percutaneous cholecystostomy, within 10 days or after 10 days, less than 2 weeks or more than 2 weeks, less than 4 weeks or more than 4 weeks, less than 8 weeks or more than 8 weeks as per literature.

**Results:** Six studies including 18640 patients were included in the final analysis. There was no difference in overall complications within or after 72 hours cholecystectomy group, but mortality and biliary complications were significantly high in the less than 72 hours group (p=0.05 and 0.0002 respectively). There was no difference in mortality, overall complication, biliary tract complications in less than 1 week versus more than 1 week and less than 10 days versus more than 10 days group. Overall complications were significantly less in the less than 2 weeks group compared to the more than 2 weeks group. There was no difference in mortality and biliary tract complications between less than 2 weeks and more than 2 weeks group. Overall complication rate (risk ratio 0.67, p <0.0001), postoperative mortality (risk ratio 0.46, p=0.003), bile duct injury (risk ratio 0.62, p=0.01) was significantly less in earlier than 4-week group. Hospital stay was not significantly different between less than 4 weeks versus more than 4 weeks group. (Mean difference= -2.74, p=0.12). Ove all complication rates were significantly more in less than 8 weeks group. (Risk ratio 1.07, p=0.01). Hospital stay was significantly less in less than 8 weeks group. (Mean difference 0.87, p=0.01).

**Conclusion:** Early cholecystectomy preferably within 4 weeks after percutaneous cholecystostomy is preferable over late cholecystectomy.

## Introduction

Acute cholecystitis is a very common presentation in surgical practice. Laparoscopic cholecystectomy is the gold standard surgical procedure to treat acute cholecystitis. Earlier it was believed that acute cholecystitis should be treated conservatively initially and then interval cholecystectomy should be offered, but now early cholecystectomy preferably within 72 hours of presentation is preferred over interval cholecystectomy. [1,2,3]

Percutaneous cholecystostomy is now emerging as a bridge to surgery to control sepsis when a patient is too sick for surgery or when a patient is unfit for surgery or has a high risk of postoperative mortality. [4] In the case of acute calculous cholecystitis, cholecystectomy is needed after a patient is stabilized with percutaneous cholecystostomy. However, the timing of subsequent cholecystectomy remains controversial, and very little data is available regarding that.

## Aim

This systematic review and meta-analysis aimed to evaluate outcomes of early versus late cholecystectomy after percutaneous cholecystectomy.

## Methods

The study was conducted according to the PRISMA 2020 statement and MOOSE guidelines. [5,6].

### Study selection

We conducted a literature search as described by Gossen et al. [7] PubMed, Cochrane Library, Embase, Google Scholar, Web of Science with keywords and “MESH” terms like “percutaneous cholecystostomy”, “Cholecystectomy”, “Timing of cholecystectomy”, “early versus late cholecystostomy”, “cholecystectomy AND cholecystostomy”, “cholecystectomy after cholecystostomy”. Two independent authors extracted the data (B.V. and H.P.) Discussions and mutual understanding resolved any disagreements.

### Statistical analysis

The meta-analysis was conducted using Review Manager 5.4. Heterogeneity was measured using Q tests and I2, and P < .10 was determined as significant, the random-effects model was used. The odds ratio (OR) was calculated for dichotomous data, and weighted mean differences (WMD) were used for continuous variables. Both differences were presented with 95% CI. For continuous variables, if data were presented with medians and ranges, then we calculated the means and standard deviations according to Hozo et al. [8]. If the study presented the median and interquartile range, the median was treated as the mean, and the interquartile ranges were calculated using 1.35 SDs, as described in the Cochrane handbook. [9]

### Risk of bias assessment

Cohort studies were assessed for bias using the Newcastle –Ottawa Scale. [10]. It was decided to assess randomized trials based on the Cochrane Handbook.[9]. However, in the final analysis, we could not find any randomized clinical trials fulfilling our inclusion criteria so the Newcastle – Ottawa Scale was used. We evaluated publication bias by funnel plots. We defined mortality as postoperative 90 days mortality. We evaluated various periods of cholecystectomy after percutaneous cholecystectomy like within 72 hours, within 7 days, within 10 days, within two weeks, within 4 weeks, or within or after 8 weeks.

### Inclusion criteria

- Studies that evaluated early vs late cholecystectomy after percutaneous cholecystostomy.
- Full-text articles.
- Conference abstracts that contain adequate information.

### Exclusion criteria

- Studies which does not contain early vs late cholecystectomy after percutaneous cholecystostomy.
- Articles where full texts were not available.
- Conference abstracts without adequate details.

## Results

Six studies including 18640 patients were included in the final analysis. [11-17]. [Figure 1]. Characteristics of the included studies are described in table 1. The risk of bias summary is shown in figure 2.

**Table 1.** Study Characteristics.

| Study | Published year | Type of study | Definition of<br>early<br>cholecystectomy<br>post<br>percutaneous<br>cholecystostomy | Total patients |
| --- | --- | --- | --- | --- |
| Han et al.[11] | 2011 | Retrospective<br>cohort | Within 72 hours | 67 |
| Jung et al.[13]. | 2015 | Retrospective<br>cohort | Within 7-10<br>days | 74 |
| Lyu et al.[14] | 2021 | Retrospective cohort | 4 weeks | 100 |
| Sakamoto et al[12]. | 2019 | Retrospective cohort | 4 weeks | 9256 |
| Alteri et al.[16] | 2019 | Retrospective cohort | 8 weeks | 2998 |
| Woodward et al.[15] | 2020 | Retrospective cohort | 8 weeks | 6145 |

**Figure 1:**
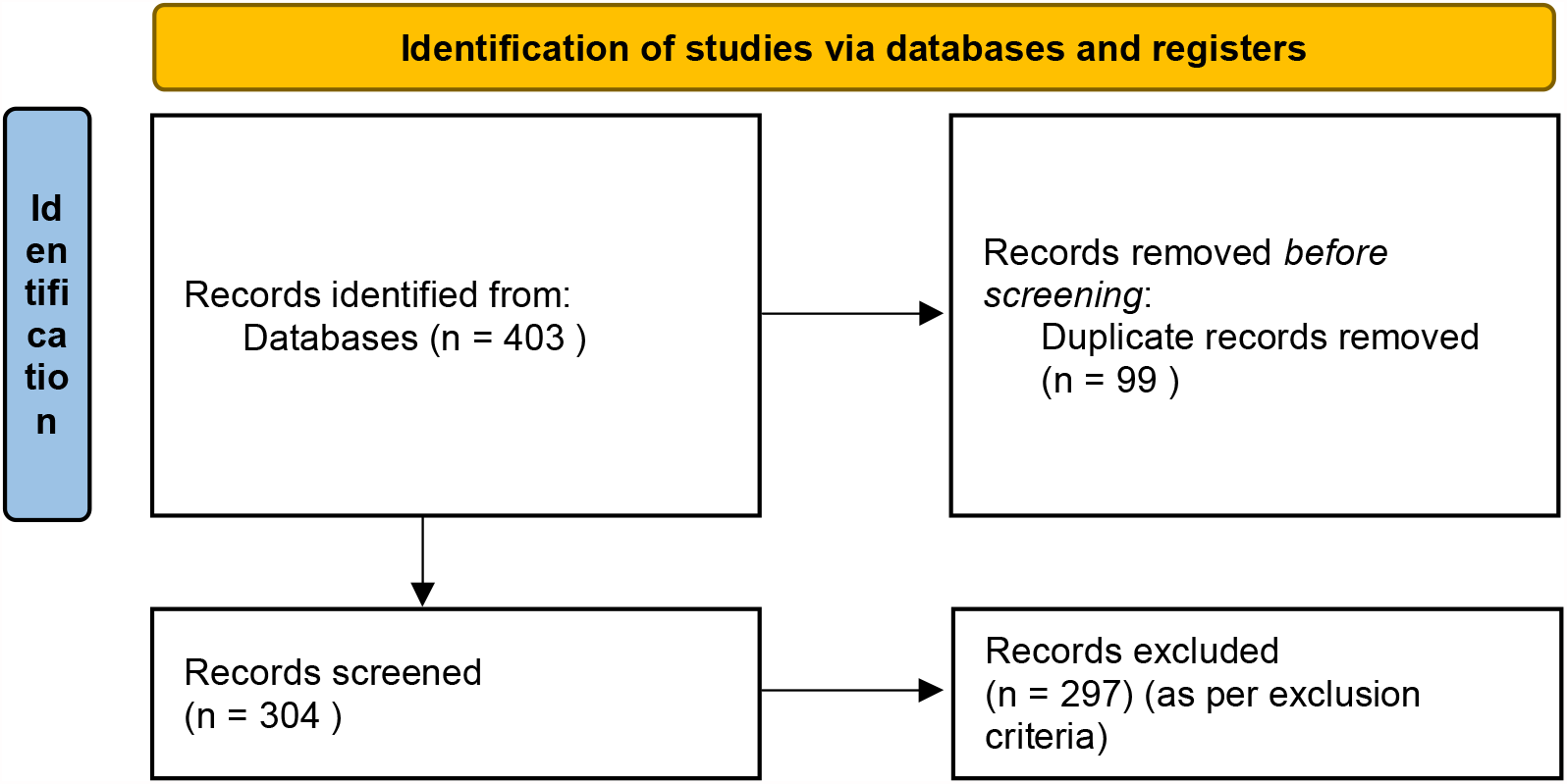
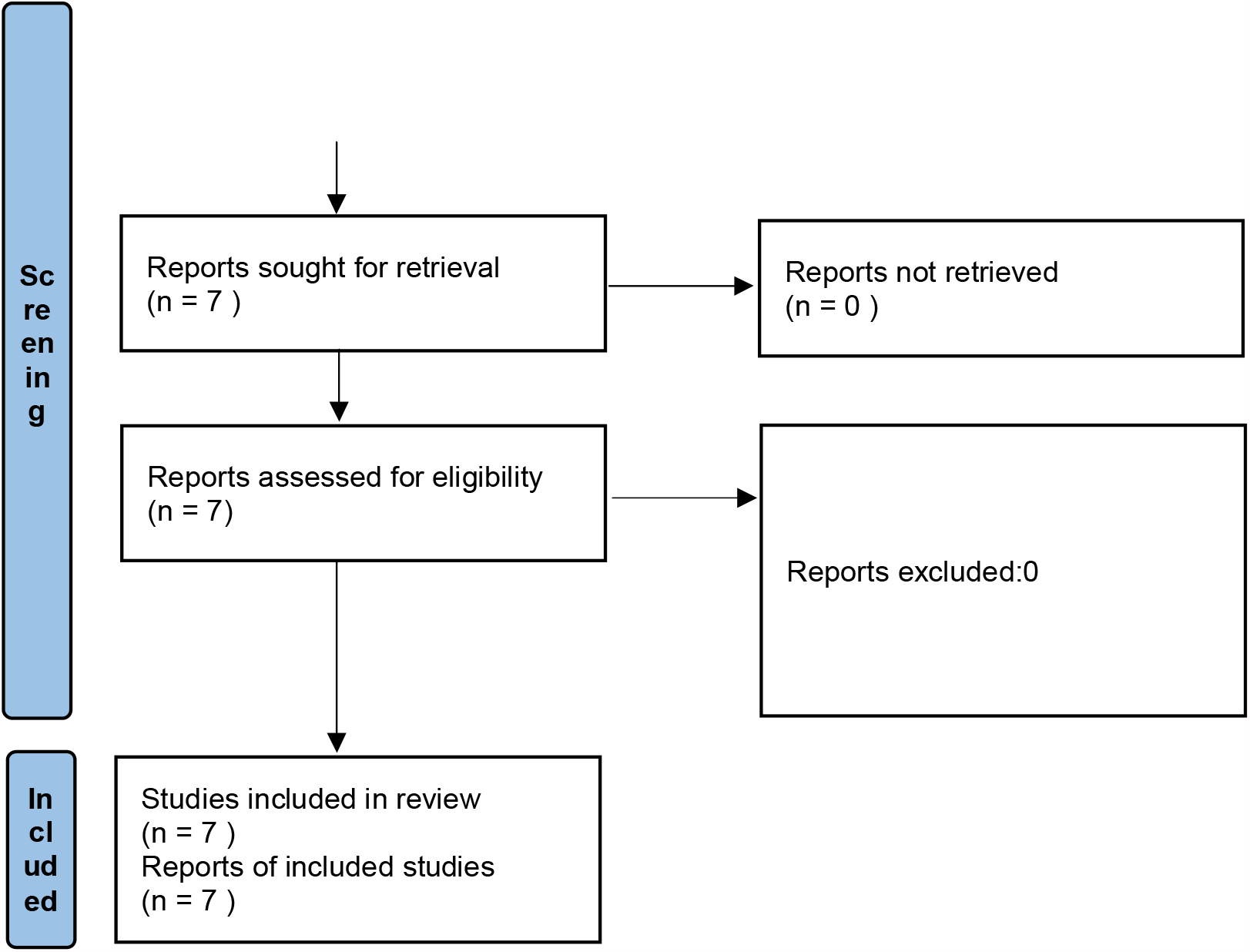
Prisma flow chart. *From:* Page MJ, McKenzie JE, Bossuyt PM, Boutron I, Hoffmann TC, Mulrow CD, et al. The PRISMA 2020 statement: an updated guideline for reporting systematic reviews. BMJ 2021;372:n71. doi: 10.1136/bmj.n71

**Figure 2.**
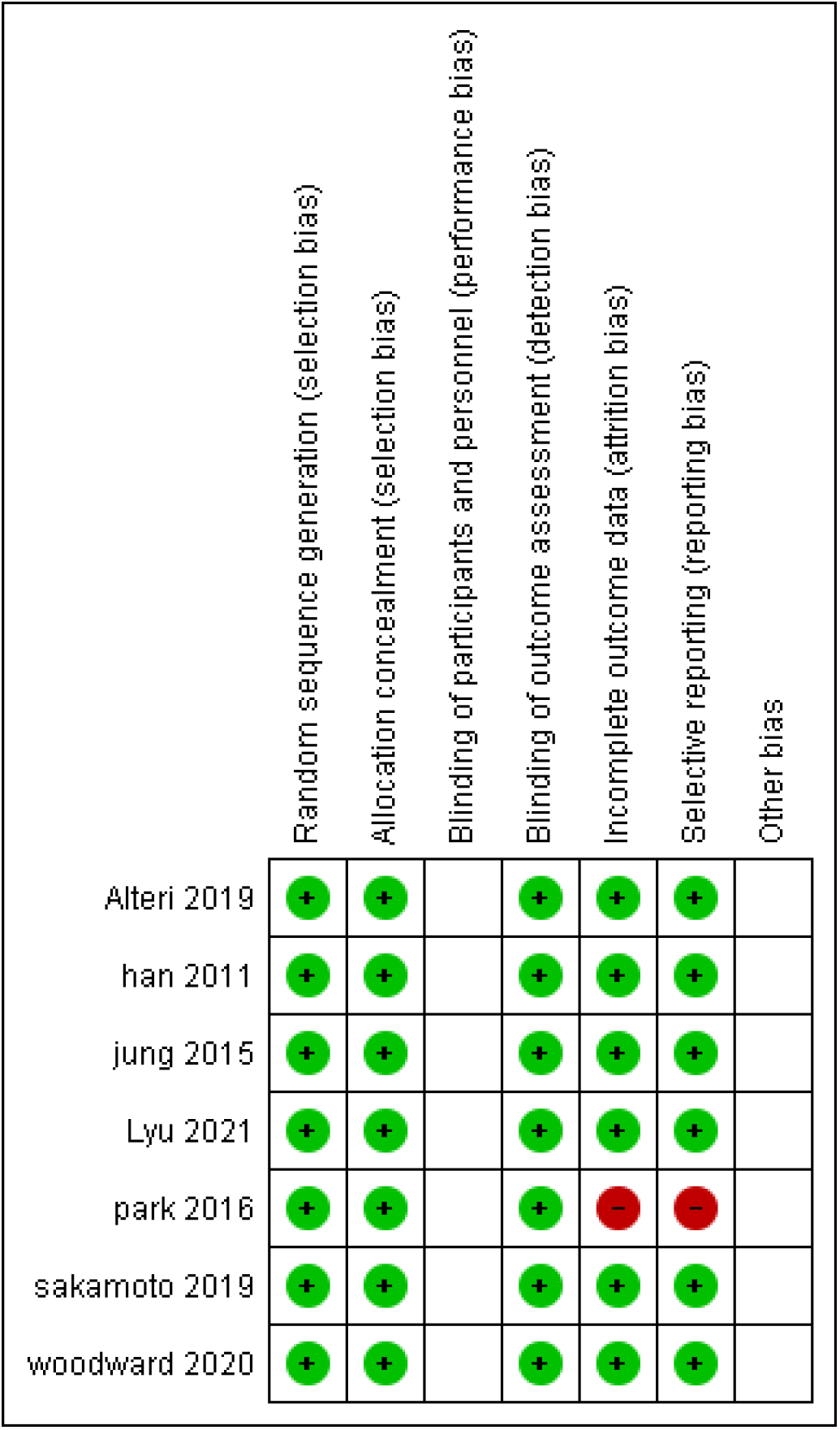
Risk of bias summary (green denotes low risk of bias, red high risk of bias and empty spaces swhos unclear bias)

### Cholecystectomy within 72 hours of percutaneous cholecystostomy: [Figure 3]

Two studies consisting of 9323 patients evaluated cholecystectomy within or after 72 hours of initial percutaneous cholecystostomy. [11,12]. There was no difference in overall complications and conversion to open between the groups. (p=0.43, p= 0.41 respectively). Biliary complications were significantly less in cholecystectomy within 72 hours. (Risk ratio 0.36, p= 0.0002). 90 days mortality was significantly higher in the early cholecystectomy group. (Risk ratio 1.8, p=0.05).

**Figure 3.**
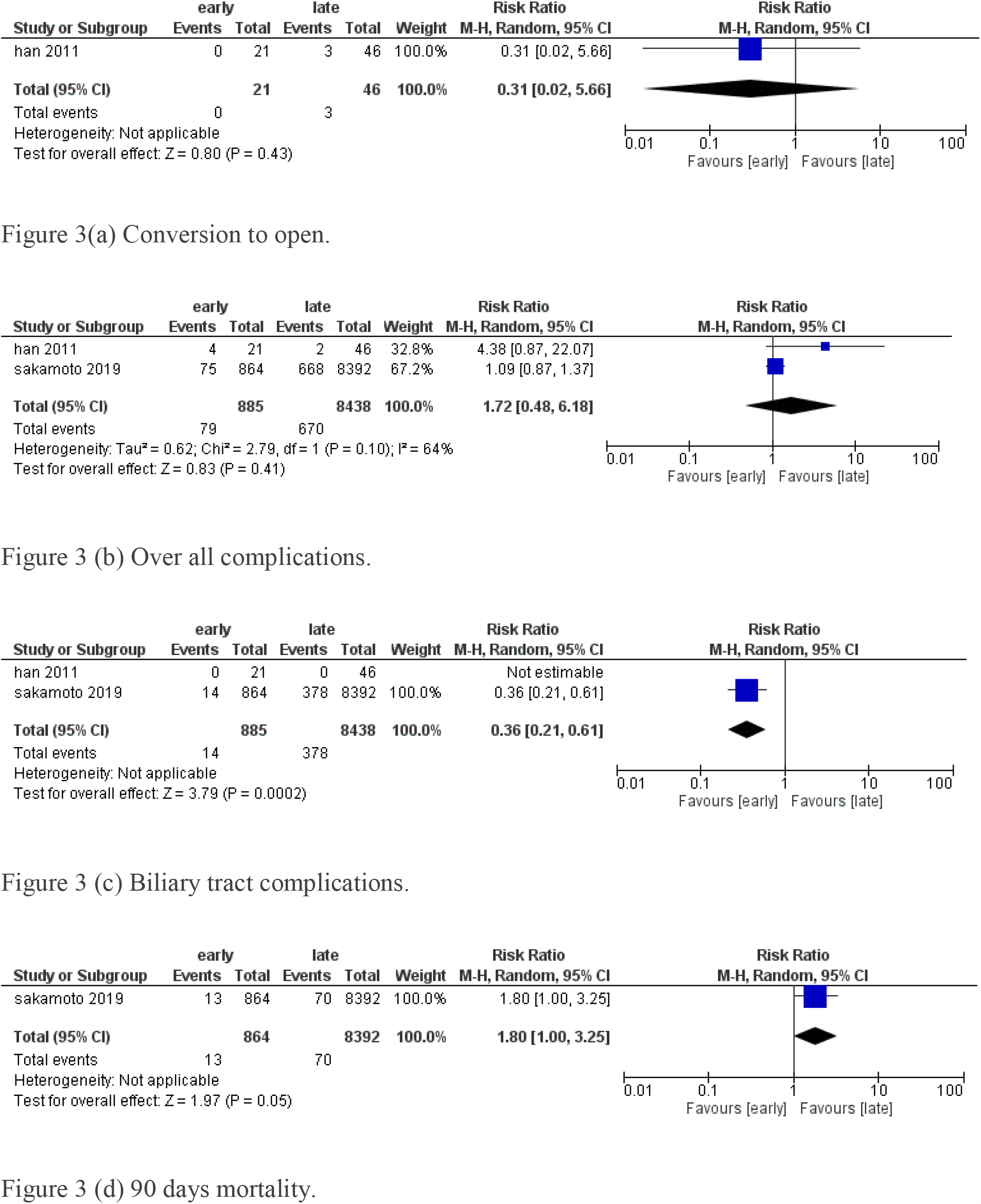
Comparisons between cholecystectomy within or after 72 hours after percutaneous cholecystostomy.

### Cholecystectomy within 1 week of percutaneous cholecystostomy: [Figure 4]

One study consisting of 9256 patients [12] compared cholecystectomy earlier than one week after percutaneous cholecystostomy to cholecystectomy after one week. There was no significant difference between overall complications, 90 days mortality, and biliary complications. (p= 0.23,0.85.0.27 respectively).

**Figure 4.**
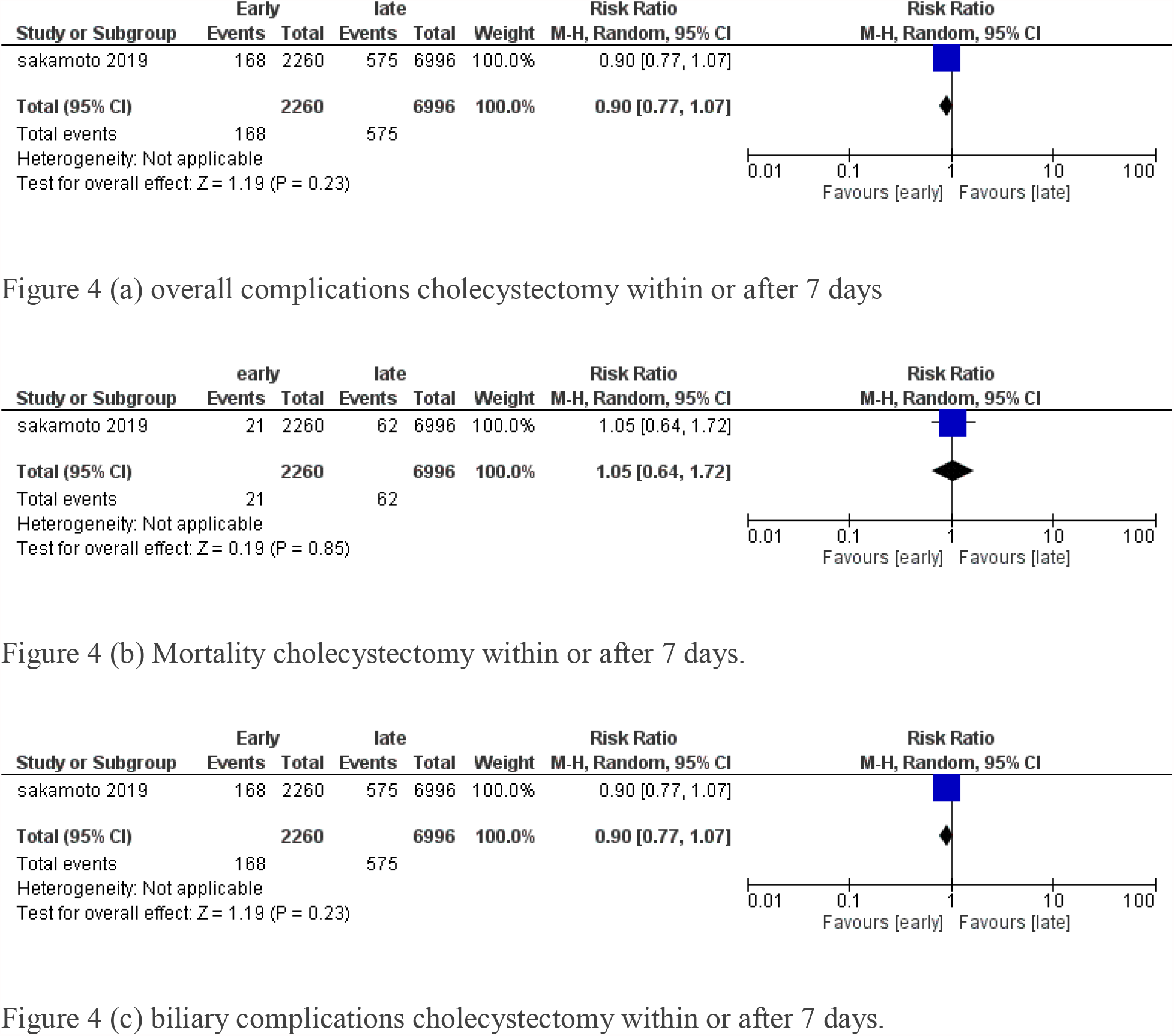
Comparisons between cholecystectomy within or after 7 days after percutaneous cholecystostomy.

### Cholecystectomy within 10 days of percutaneous cholecystostomy: [Figure 5]

One study with 74 patients [13] evaluated cholecystectomy within or after 10 days of percutaneous cholecystostomy. There was no difference in conversion to open, overall complications, and bile duct injury between cholecystectomy within or after 10 days. (p=0.43,0.36.0.36 respectively).

**Figure 5.**
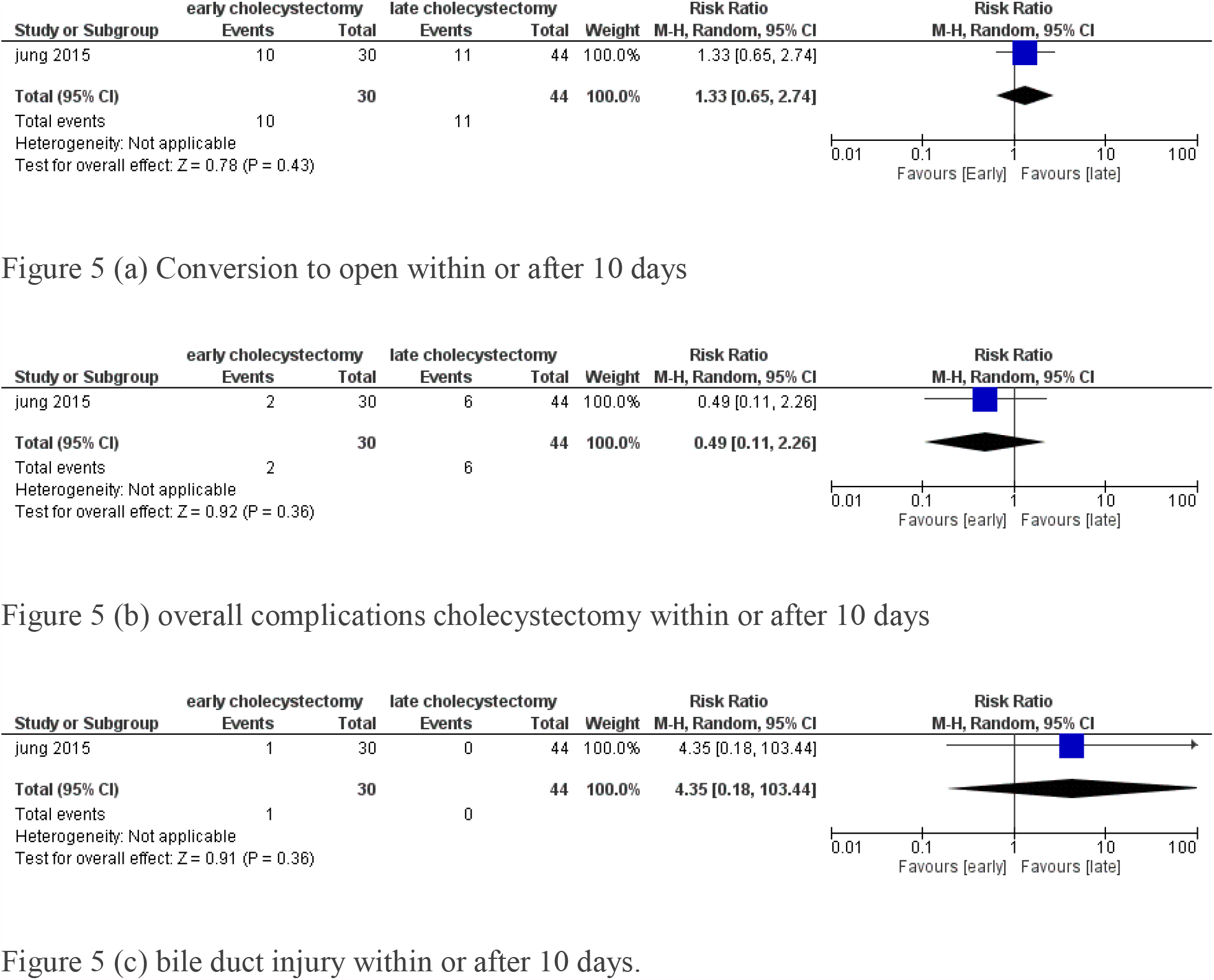
Comparisons between cholecystectomy within or after 10 days after percutaneous cholecystostomy.

### Cholecystectomy within 2 weeks of percutaneous cholecystostomy: [Figure 6]

One study consisting of 9256 patients [12] compared cholecystectomy earlier than two weeks after percutaneous cholecystostomy to cholecystectomy after two weeks. Overall complications were significantly less in cholecystectomy earlier than two weeks. (p=0.02, Risk ratio 0.85). There was no difference in 90 days mortality and biliary complications. (p=0.48,0.34 respectively)

**Figure 6.**
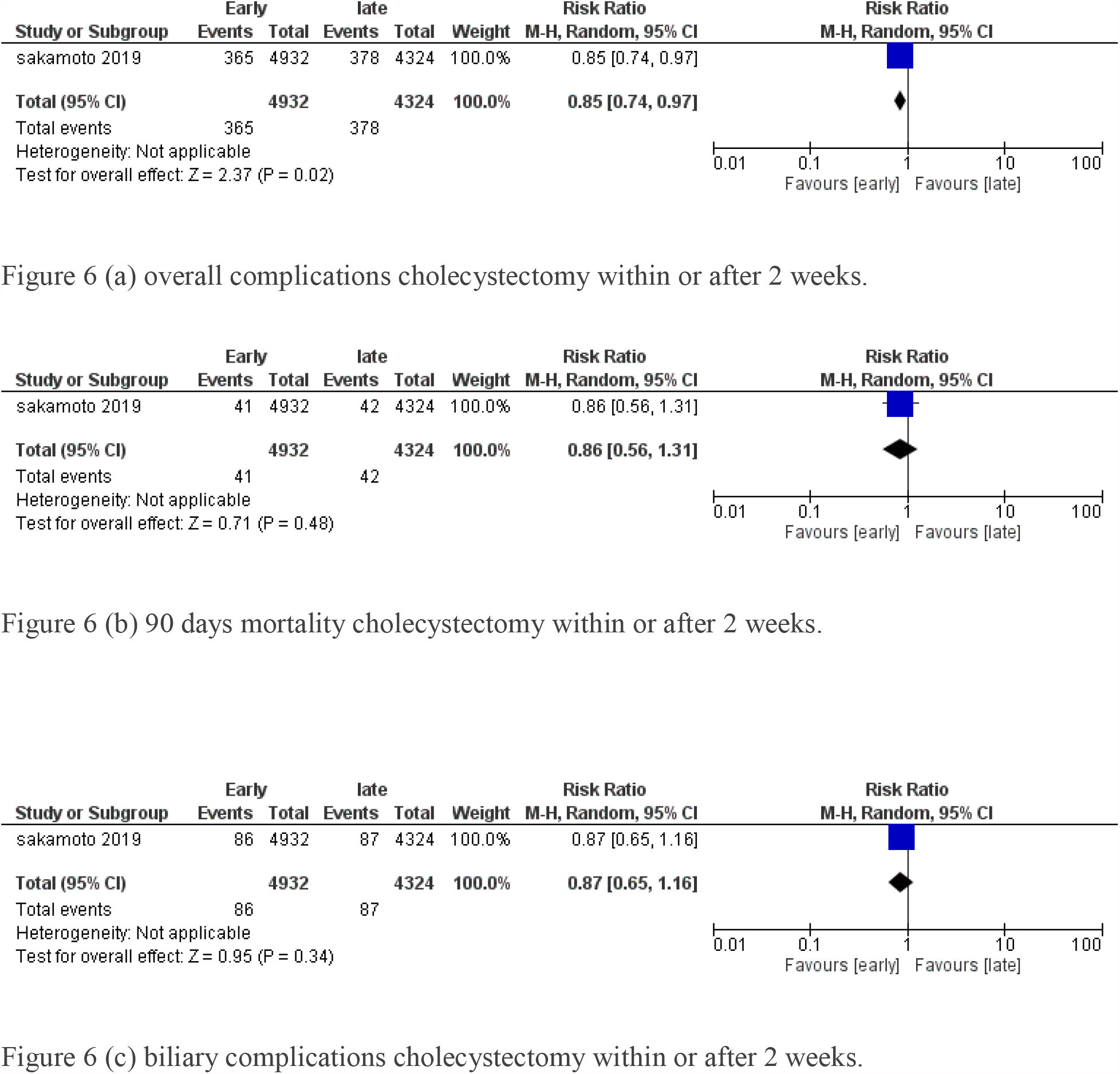
Comparisons between cholecystectomy within or after 2 weeks after percutaneous cholecystostomy.

### Cholecystectomy within 4 weeks of percutaneous cholecystostomy: [Figure 7]

Two studies consisting of 9356 patients evaluated cholecystectomy within or after 4 weeks of initial percutaneous cholecystostomy. [12,14]. There was no difference in conversion to open in both groups. (p=0.13). Overall complications were significantly less in the within 4 weeks cholecystectomy group. (risk ratio 0.67, p <0.00001), 90 days mortality was also significantly less in cholecystectomy within 4 weeks. (Risk ratio 0.46, p=0.003). Biliary tract complications were significantly low in the early cholecystectomy group. (Risk ratio 0.62, p=0.01). There was no difference in hospital stay between the two groups. (p=0.12)

**Figure 7.**
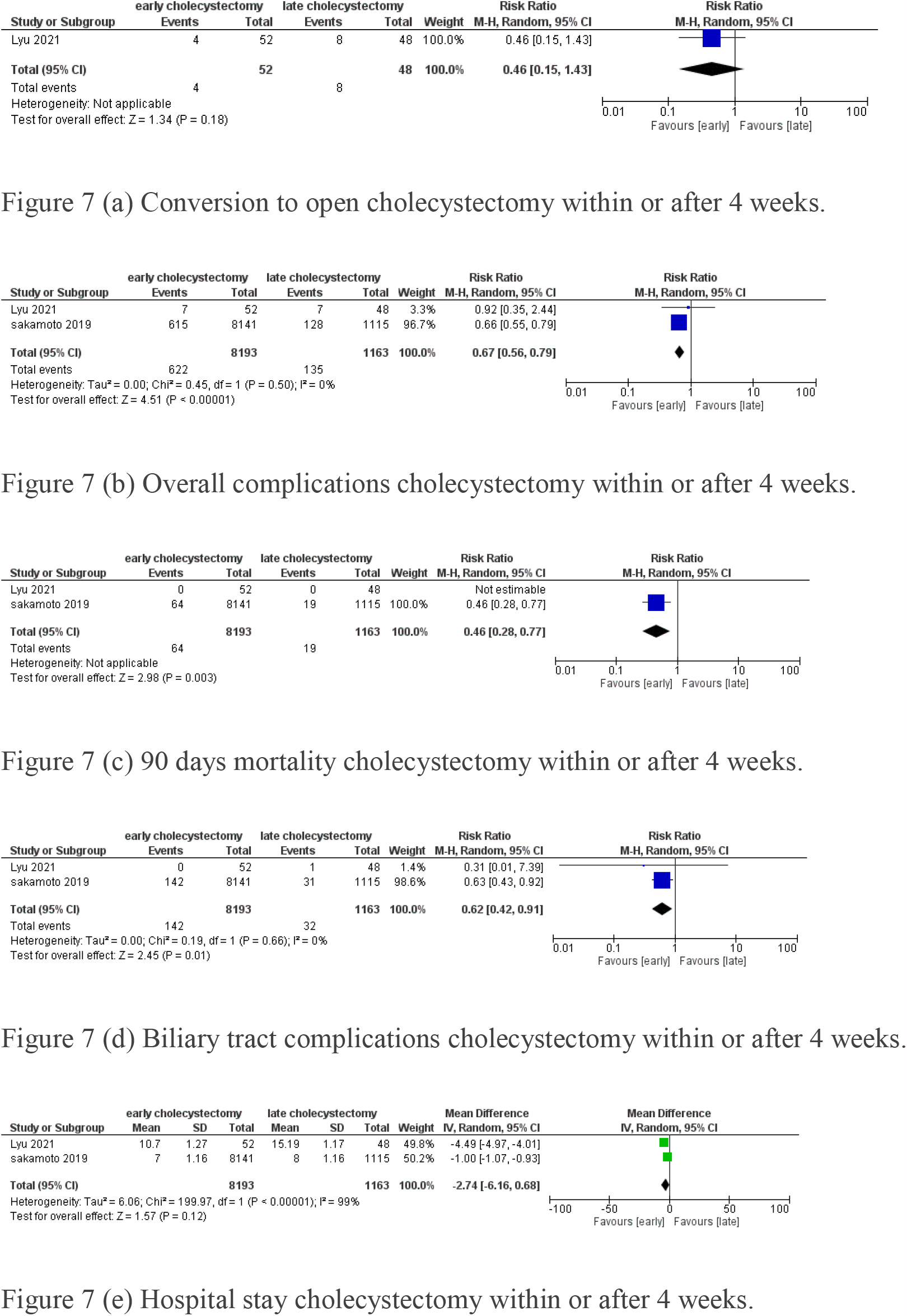
Comparisons between cholecystectomy within or after 4 weeks after percutaneous cholecystostomy.

### Cholecystectomy within 8 weeks of percutaneous cholecystostomy: [Figure 8]

Two studies consisting of 9143 patients evaluated cholecystectomy within or after 4 weeks of initial percutaneous cholecystostomy. [15,16]. There was no difference in conversion to open in both groups. (p=0.77). Overall complications were significantly more in within 8 weeks cholecystectomy group. (risk ratio 1.07, p =0.01). There was no difference in 90 days mortality between the groups. (p=0.57). Hospital stay was significantly more in cholecystectomy less than 8 weeks group. (mean difference 0.87, p=0.01). There was no difference in risk of subtotal cholecystectomy between groups. (p=0.75).

**Figure 8.**
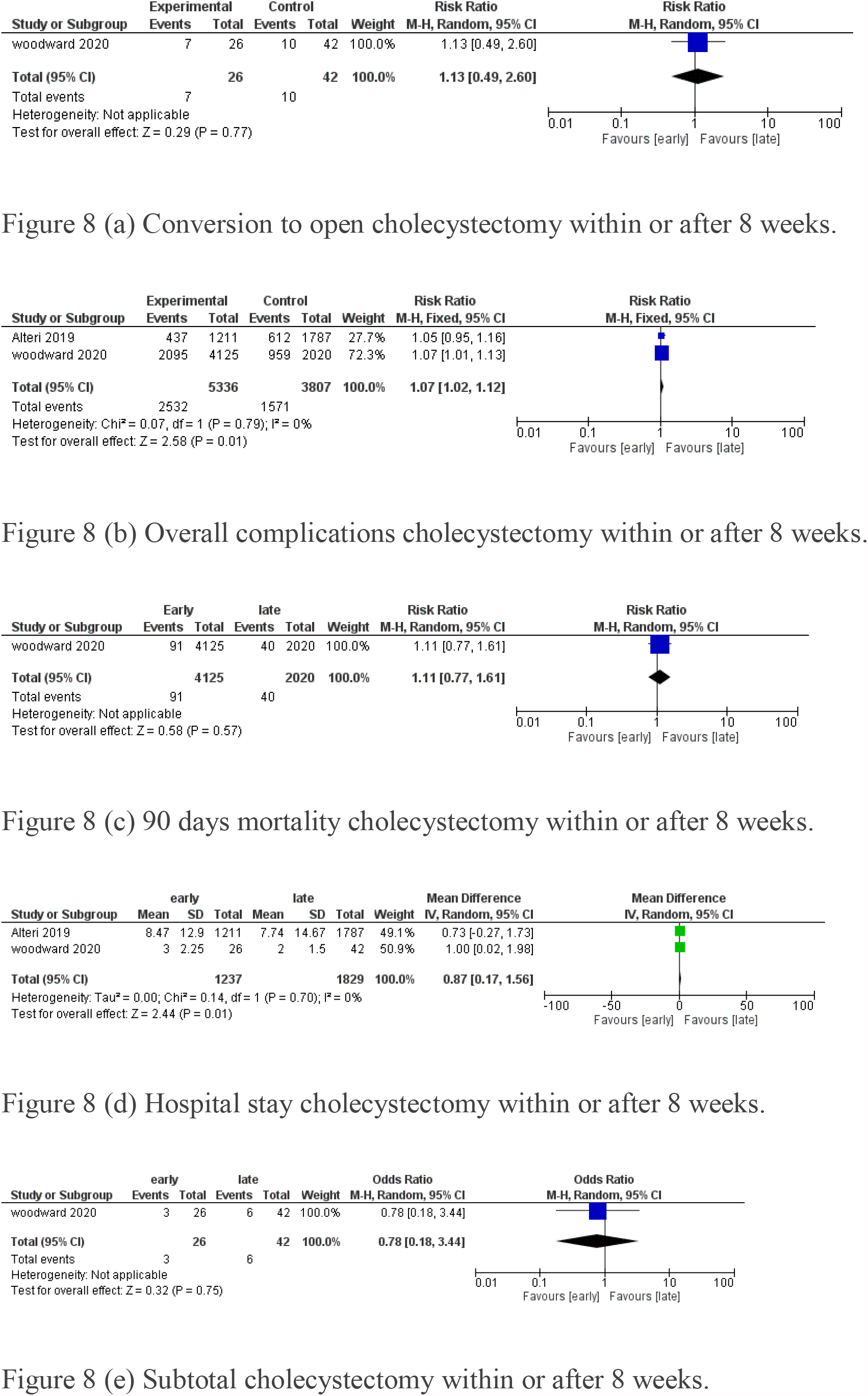
Comparisons between cholecystectomy within or after 8 weeks after percutaneous cholecystostomy.

Heterogeneity was not significant in the majority of analysis as shown in forest plots, in some of the heterogeneity analysis was not possible as very few studies were there. Publication bias was not significant.

## Discussion

Percutaneous cholecystostomy is increasingly being used for definitive treatment of acalculous cholecystitis and as a bridge to surgery in case of calculous cholecystitis in a patient with a high risk of surgery due to comorbidities or sometimes ongoing septic shock. [16-20]. There is ongoing controversy about the optimal time of cholecystectomy after percutaneous cholecystostomy in the case of calculous cholecystectomy.

In this systematic review and meta-analysis, we tried to evaluate the optimal timing of cholecystectomy after percutaneous cholecystostomy in the case of calculous cholecystitis.. We evaluated conversion to open, overall complications after cholecystectomy, biliary complications, 90-day mortality, hospital stay after cholecystectomy performed in various time frames after cholecystostomy based on available literature.

After a systematic search described in methodology and removing duplicates, we found six articles [11-17], which evaluated the timing of cholecystectomy after percutaneous cholecystostomy. There was a lack of uniformity about the timing of cholecystectomy after percutaneous cholecystostomy in the available literature, so we evaluated different time frames in our meta-analysis.

As per findings of the meta-analysis, there was no increase in conversion to open, and overall complications of cholecystectomy are performed within 72 hours of percutaneous cholecystostomy, biliary complications were lesser if cholecystectomy is performed within 72 hours of percutaneous cholecystostomy. However, 90 days mortality was slightly higher (p=0.05) if cholecystectomy is performed within 72 hours but we are not sure this is due to cholecystectomy or due to underlying conditions. [figure 3].

We found no increase in mortality, overall complications, biliary complications, and conversion to open if cholecystectomy is performed within 7 and 10 days after percutaneous cholecystostomy. [figure 4,5]. Only one study [12] evaluated cholecystectomy within or after two weeks of percutaneous cholecystostomy, which also showed early cholecystectomy reduced overall complications without increasing mortality and biliary complications. [figure 6].

Maximum benefit is observed as per our meta-analysis when cholecystectomy is performed within 4 weeks of percutaneous cholecystostomy. It reduced overall complications, biliary complications, 90 days mortality, hospital stay without increasing conversion to open cholecystectomy. [figure 7].

In studies comparing cholecystectomy within or after 8 weeks of percutaneous cholecystostomy [15,16], overall complications and hospital stay were significantly more in cholecystectomy in less than 8 weeks compared to cholecystectomy in more than 8 weeks. There was no difference in conversion to open, 90-day mortality and need for subtotal cholecystectomy between the groups.

Based, on these findings, it seems that less than 4 weeks is the ideal time for cholecystectomy after percutaneous cholecystostomy for calculus cholecystitis.

There were certain limitations in our meta-analysis, first, we could include only a limited number of studies as there is still a limited number of studies done in this research question. Three studies [12,15,16] had the majority of patients and other studies included a very limited number of patients and hence results can be skewed by the weight given to those three studies during various analyses. Also, the time duration of cholecystectomy after cholecystostomy varied widely across the studies. So, we had to analyze various time frames.

To our knowledge, this is the first meta-analysis evaluating the optimal time duration of cholecystectomy after percutaneous cholecystostomy. Also, analysis across various time frames confirmed that earlier cholecystectomy is not harmful as it was thought. The ideal time duration from our analysis seems earlier than 4 weeks, most probably 1-4 weeks. The most favorable findings are fewer overall and biliary complications in early cholecystectomy without increased need to convert to open cholecystectomy. However, still, studies with a larger number of patients or randomized control trials are needed to confirm our findings.

In conclusion, early cholecystectomy preferably within 4 weeks after cholecystostomy is safe and probably beneficial to the patients. However, further studies are still needed to confirm the findings of this meta-analysis.

## Data Availability

Will be made available on demand

## Notes

Conflict of interest: None

Funding disclosure: none

### Competing Interest Statement

The authors have declared no competing interest.

### Funding Statement

No funding obtained

